# Sociodemographic inequalities and excess non-COVID-19 mortality during the COVID-19 pandemic: A data-driven analysis of 1,069,174 death certificates in Mexico

**DOI:** 10.1101/2022.05.12.22274973

**Authors:** Neftali Eduardo Antonio-Villa, Carlos A. Fermín-Martínez, José Manuel Aburto, Luisa Fernández-Chirino, Daniel Ramírez-García, Julio Pisanty-Alatorre, Armando González-Díaz, Arsenio Vargas-Vázquez, Jacqueline A. Seiglie, Simón Barquera, Luis Miguel Gutiérrez-Robledo, Omar Yaxmehen Bello-Chavolla

## Abstract

**BACKGROUND:** In 2020, Mexico experienced one of the highest rates of excess mortality globally. However, the extent to which non-COVID deaths contributed to excess mortality, its regional characterization, and the association between municipal-and individual-level sociodemographic inequality has not been characterized.

**METHODS:** We conducted a retrospective municipal an individual-level study using death certificate data in Mexico from 2016-2020. We analyzed mortality related to COVID-19 and to non-COVID-19 causes using ICD-10 codes to identify cause-specific mortality. Excess mortality was estimated as the increase in deaths in 2020 compared to the average of 2016-2019, disaggregated by primary cause of death, death setting (in-hospital and out-of-hospital) and geographical location. We evaluated correlates of non-COVID-19 mortality at the individual level using mixed effects logistic regression and correlates of non-COVID-19 excess mortality in 2020 at the municipal level using negative binomial regression.

**RESULTS:** We identified 1,069,174 deaths in 2020 (833.5 per 100,000 inhabitants), which was 49% higher compared to the 2016-2019 average (557.38 per 100,000 inhabitants). Overall excess mortality (276.11 deaths per 100,000 inhabitants) was attributable in 76.1% to COVID-19; however, non-COVID-19 causes comprised one-fifth of excess deaths. COVID-19 deaths occurred primarily in-hospital, while excess non-COVID-19 deaths decreased in this setting and increased out-of-hospital. Excess non-COVID-19 mortality displayed geographical heterogeneity linked to sociodemographic inequalities with clustering in states in southern Mexico. Municipal-level predictors of non-COVID-19 excess mortality included levels of social security coverage, higher rates of COVID-19 hospitalization, and social marginalization. At the individual level, lower educational attainment, blue collar workers, and lack of medical care assistance were associated with non-COVID-19 mortality during 2020.

**CONCLUSION:** Non-COVID-19 causes of death, largely chronic cardiometabolic conditions, comprised up to one-fifth of excess deaths in Mexico during 2020. Non-COVID-19 excess deaths occurred disproportionately out-of-hospital and were associated with both individual-and municipal-level sociodemographic inequalities. These findings should prompt an urgent call to action to improve healthcare coverage and access to reduce health and sociodemographic inequalities in Mexico to reduce preventable mortality in situations which increase the stress of healthcare systems, including the ongoing COVID-19 pandemic.

## INTRODUCTION

Over 6 million deaths attributable to COVID-19 have been reported globally since the onset of the pandemic in early 2020 (1). Beyond this devastating death toll, there is increasing recognition of the widespread disruption of the pandemic on healthcare services, with a far-reaching impact on the care of non-COVID-19 conditions (2). Excess mortality has been proposed as a key indicator which captures both deaths caused by COVID-19 and indirect deaths attributed to the pandemic more broadly, due to interruption in routine care of chronic conditions (3,4). Notably, many low-and-middle-income countries (LMICs), particularly in Latin America, were vulnerable to the direct and indirect effects of the COVID-19 pandemic due to chronic underinvestment in public healthcare (5). Though several reports have estimated that rates of excess mortality were disproportionately higher in LMICs following the onset of the COVID-19 pandemic, there is limited insight regarding non-COVID-19 deaths and their contribution to the reported rates of excess mortality in Latin America (6).

Mexico is of particular interest given that it ranks as one of the countries with the highest rates of excess mortality in the Latin American region following onset of the COVID-19 pandemic. (7). A confluence of health and sociodemographic inequalities that predated the COVID-19 pandemic, a high burden of chronic cardiometabolic conditions, and a fragmented healthcare system all contributed to a high and disproportionate burden of excess mortality among marginalized communities (5,8). A descriptive assessment performed in Mexico showed that chronic cardiometabolic conditions, which are highly prevalent among communities of low socioeconomic status, were the main causes of death independently of registered COVID-19 deaths in Mexico during 2020 (9). However, whether hospital saturation had ripple effects on out-of-hospital excess mortality, particularly for highly prevalent chronic health conditions across different vulnerable regions, has not yet been characterized. Hence, there is a need to comprehensively assess the extent to which individual and municipal sociodemographic inequalities impacted excess mortality to further guide health policies to strengthen existing systems and mitigate ongoing health disparities. In this study, we sought to: 1) estimate the age-adjusted rates of cause-specific excess mortality due to COVID-19 and non-COVID-19 deaths in 2020 compared to the 2016-2019 period, stratified by in-hospital and out-of-hospital setting; 2) evaluate the geographical distribution of cause-specific excess mortality in Mexico in 2020; and 3) characterize the association between municipal-and individual-level sociodemographic inequality measures and non-COVID-19-related excess mortality.

## METHODS

### Study design and data sources

We conducted a retrospective municipal and individual-level study using national mortality records from 2016-2019 compared to 2020. Death certificate records of individuals living in Mexico were collected by the National Institute of Statistics and Geography (INEGI). Briefly, INEGI generates annual mortality statistics from death certificates and vital sociodemographic characteristics issued by the Ministry of Health, which includes the primary cause of death in accordance with the 10^th^ version of the International Statistical Classification of Diseases and Related Health Problems (ICD-10, (10)). Complete methodology of the death certification process, validation, and collected variables are available in **Supplementary Material**.

### Variables and definitions

#### Outcome variables

Cause-specific excess mortality was centered on two primary outcomes: deaths due to COVID-19 and deaths related to non-COVID-19 causes. Overall excess mortality was the sum of excess mortality due to non-COVID-19 causes and all registered COVID-19 deaths.

##### I. COVID-19 deaths

Deaths attributable to COVID-19 were defined under the following ICD-10 codes: U071 (identified SARS-CoV-2), U072 (suspected SARS-CoV-2), and deaths after April-2020 classified as J00-J99 (respiratory deaths). This aggregation of COVID-19 deaths considers inadequate registration of COVID-19 cases across 2020, as there is an unknown number of deaths which could have been classified with unspecified respiratory diseases in the early stages of the pandemic due to limited SARS-CoV-2 testing capacity in Mexico (11).

##### II. Non-COVID-19 cause-specific mortality

All other causes of death were classified as non-COVID-19 related deaths and were coded using the 2020 Mexican list of mortality, which comprises 436 specific causes of death (12). To simplify result presentation, we only display the first 10 cause-specific deaths in the main results, with the full list provided in **Supplementary material**.

#### Excess Mortality Estimation

According to the approach proposed by Karlinsky and Kobak, we estimated excess mortality as the difference between average deaths during the 2016-2019 period compared to deaths registered during 2020 (13). We used average deaths for two reasons: 1) the use of average deaths is a simple approach proven to be a reliable assessment based on sensitivity analyses estimations, and 2) given that we are estimating 436 specific causes of death, predictive methods based on generalized linear models may overestimate the standard error for low-frequency causes (9). Excess deaths were standardized to age-adjusted rates per 100,000 population with age structures by state, municipalities, and regions per 5-year increments using population projections provided by the National Population Council (CONAPO). Percent increase in 2020 compared to 2016-2019 was also used as a proxy of excess mortality.

#### Stratification by setting of death

We hypothesized that the COVID-19 pandemic posed a significant burden on in-hospital care, which may have influenced increases in excess mortality, particularly for non-COVID-19 related deaths. To evaluate this, we stratified excess mortality according to whether the death occurred out-of-hospital or in-hospital, as registered in death certificates. Out-of-hospital deaths were defined accordingly if the death was not registered in a hospital setting or if they were coded as occurring at the deceased person’s home or elsewhere (i.e., in the streets in some instances). Deaths with an unspecified setting were excluded across all the analysis.

#### Marginalization Index

To quantify the impact of municipal socio-demographic inequalities on excess mortality, we used the 2020 municipal social lag index (SLI) estimated by the National Council for Evaluation of Social Development Policy (CONEVAL, (14)). Since we intended to evaluate social inequalities independently from urbanization and centralized health services, we used residuals of linearly regressed mean urban population density (MUPD) and hospital beds per 100,000 inhabitants using data extracted from CONEVAL to fit an adjusted municipal SLI (aSLI). We then categorized municipalities into four aSLI categories (Low-aSLI, Moderate-aSLI, High-aSLI, and Very-High aSLI) based on the Dalenius & Hodges method (**Supplementary Material**).

#### Municipal healthcare determinants of excess mortality

The percentage of the population without healthcare coverage and the hospital occupancy due to COVID-19 inpatients were evaluated as municipal healthcare correlates of excess mortality. Healthcare coverage was obtained from 2020 CONEVAL estimations. To estimate a surrogate of hospital occupancy, we used the number of hospitalizations with confirmed COVID-19 from the SINAVE dataset collected by the General Directorate of Epidemiology (*DGE*) of the Mexican Ministry of Health, which includes reports of daily updated suspected COVID-19 cases (15). Complete methodology, the protocol of testing, and the variables included are available in **Supplementary Material**.

### Statistical analysis

To visualize differences in deaths over time in 2020 compared with the 2016-2019 period, we first plotted excess mortality per 100,000 inhabitants by month of occurrence, stratified by COVID-19 and non-COVID-19 causes. We then disaggregated excess mortality rates due to COVID-19 or non-COVID-19 causes by state and municipality. Next, to visualize whether the proportion of age-adjusted excess mortality in each municipality increased due to COVID-19 or non-COVID-19 causes we used choropleth maps classified using the quantile method with the *biscale* R package. We further visualized the relationship between excess mortality and aSLI using the same method. The median value for the estimated age-adjusted excess mortality and the aSLI were considered as the cut-off threshold.

Next, we evaluated the impact of municipal characteristics on increased risk of age-adjusted excess mortality, using negative binomial regression models to obtain incidence rate ratios (IRRs). Models were adjusted for sex, education, access to medical assistance, and urbanization (to adjust for residual covariance). We also calculated the ratio of out-of-hospital to in-hospital deaths, which was also adjusted for the above outlined covariates (16). IRRs were plotted using the *jtools* R Package. To evaluate the individual-level probability of death attributable to non-COVID-19 causes as compared to COVID-19, we fitted hierarchical random-effects logistic regression models, which included individual and municipal-level variables. Individual-level variables included sex, education, self-reported indigenous identity, work occupation, access to medical assistance prior to death, and social security coverage. Municipal-level variables included living in municipalities with low-hospital bed occupancy (<1 bed per 100,000 inhabitants), and municipal aSLI categories. For this model, we used the municipality of death occurrence as a random intercept to account for intermunicipal variability in death registration in the model and to establish a hierarchical relationship between individual and municipal-level variables. All analyses were performed using R software (Version 4.1.2).

## RESULTS

### Overall and cause-specific excess mortality in Mexico during 2020

We identified 1,069,174 deaths in Mexico during 2020, which was higher compared to 698,080 average deaths in 2016-2019. We estimated an age-adjusted mortality rate of 833.49 deaths per 100,000 inhabitants for 2020, with an estimated age-adjusted excess mortality of 276.11 deaths per 100,000 inhabitants; this represents a 49% increase in mortality compared with the average age-adjusted mortality rates of 2016-2019 (557.38 per 100,000 inhabitants). The highest peaks of excess mortality during 2020 were recorded during the May to June period. Approximately 76.1% of excess deaths were attributable to confirmed or suspected COVID-19, whilst 23.9% were attributable to non-COVID-19 causes. The main contributors of excess mortality were suspected or confirmed COVID-19 deaths (199.26 per 100,000 inhabitants). The five leading causes of non-COVID-19 excess mortality were acute myocardial infarction (44.32 deaths per 100,000 inhabitants), type 2 diabetes (34.36 deaths per 100,000 inhabitants), hypertensive heart disease (3.12 deaths per 100,000 inhabitants), essential arterial hypertension (3.02 deaths per 100,000 inhabitants) and violent assaults (1.77 deaths per 100,000 inhabitants). Notably, all excess deaths were recorded after April 2020, with a steep increase after this period for COVID-19, acute myocardial infarction, and type 2 diabetes related deaths (**Figure 1**).

**Figure 1:**
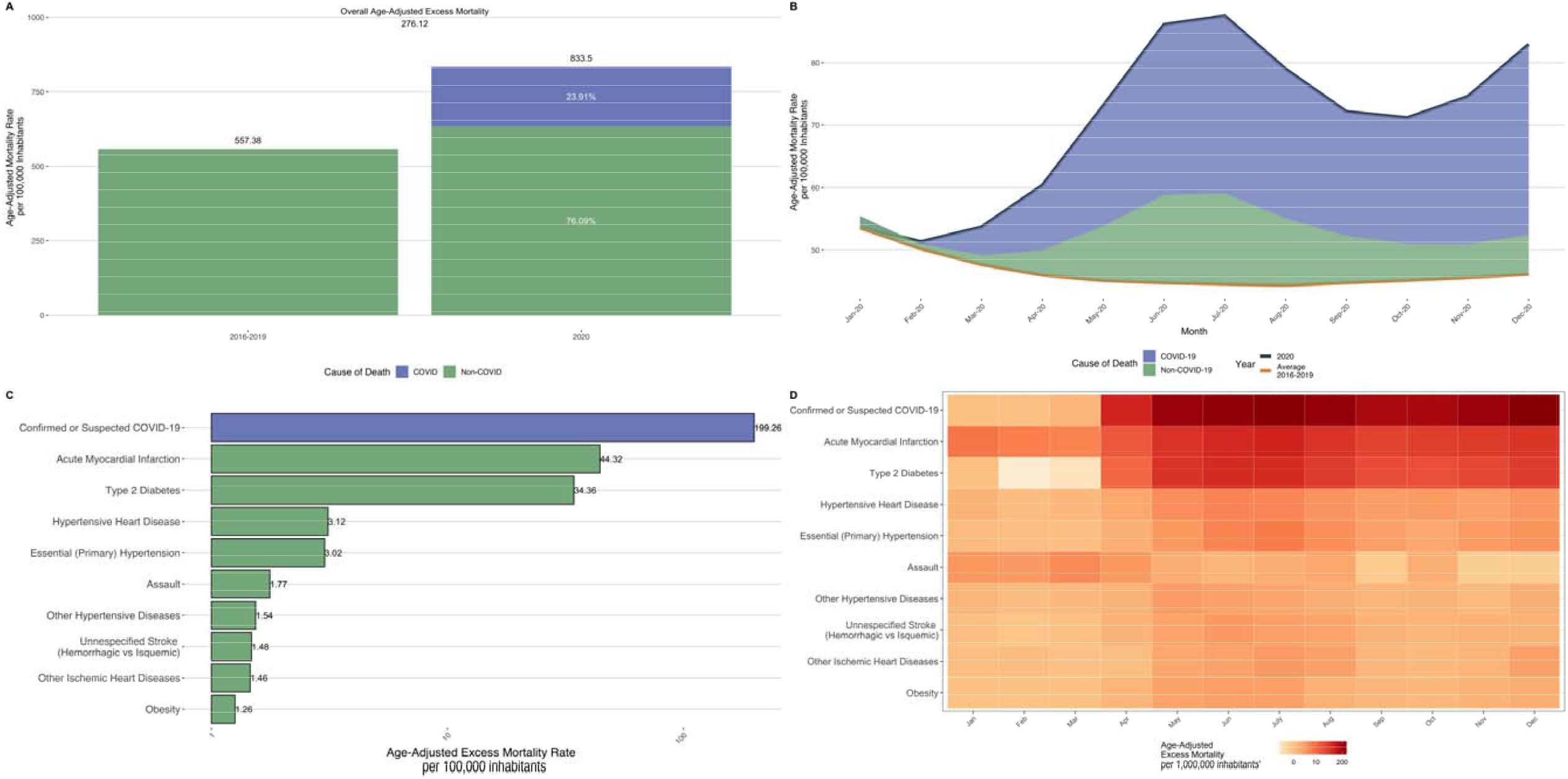
Age-adjusted mortality rates in 2020 compared by the average period of 2016-2019 (A). Distribution of excess mortality in Mexico stratified by deaths associated with COVID-19 and non-COVID-19 for the year 2020 (B). Main 10 causes of age-adjusted excess mortality in México (C) and stratified by month of occurrence (D).

### Excess mortality according to in-hospital vs. out-of-hospital death

Stratifying by the setting of death, we estimated an in-hospital excess mortality rate of 110.96 deaths per 100,000 inhabitants and an out-of-hospital excess mortality rate of 158.10 deaths per 100,000 inhabitants; this represents an increase of 44.4% and 53.2% of in-hospital and out-of-hospital deaths, respectively, compared to the average of 2016-2019. When stratified by the specific cause of death, we observed that excess in-hospital mortality rates were primarily attributable to COVID-19 deaths, while there was a decrease for in-hospital non-COVID-19 mortality rates after March 2020. Furthermore, we observed that 92.21% of all out-of-hospital excess mortality was attributable to non-COVID-19 causes, while only 6.79% were attributable to COVID-19 deaths. Regarding COVID-19 excess deaths, we observed that these causes had a higher burden of deaths in in-hospital locations compared with out-of-hospital settings, while most non-COVID-19 deaths occurred mainly out-of-hospital. We observed that acute myocardial infarction and type 2 diabetes were causes of death that decreased in-hospital, but sharply increased out-of-hospital after April-2020 (**Figure 2**).

**Figure 2:**
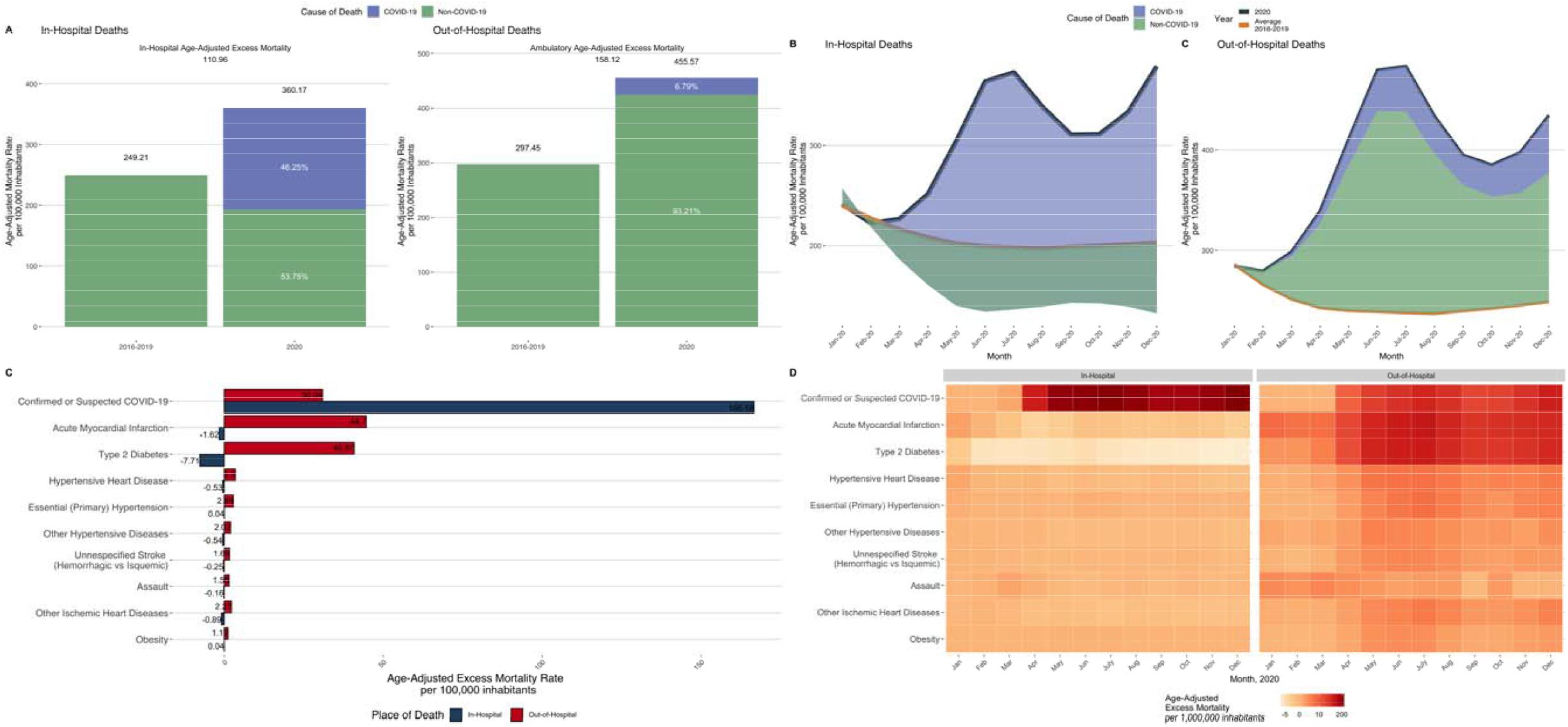
Stratification of in-hospital and out-of-hospital age-adjusted mortality rates in 2020 compared by the average period of 2016-2019 (A). Distribution of excess mortality in Mexico stratified death setting and by deaths associated with COVID-19 and non-COVID-19 for the year 2020 (B). Main 10 causes of age-adjusted excess mortality in México stratified by place of occurrence (C) and by month and place of occurrence of death (D).

### Regional state and municipal-level heterogeneity in COVID-19 and non-COVID-19 excess mortality

When COVID-19 and non-COVID-19 excess mortality was stratified at the state level, we observed that Mexico City, Baja California, and Chihuahua displayed the highest COVID-19 age-adjusted excess mortality rate, while Chihuahua, Mexico City, and Chiapas had the highest non-COVID-19 excess mortality rates. Moreover, we observed an unequal distribution of non-COVID-19 age-adjusted excess mortality at the state level, where Chiapas, Oaxaca, and Michoacán, which are in southern Mexico, displayed higher rates for non-COVID-19 causes (**Supplementary Figure 1**). Further stratification revealed a geographical aggregation of non-COVID-19 deaths caused by acute myocardial infarction, type 2 diabetes, essential arterial hypertension, and unspecified strokes conglomerated in the southern states of Mexico (**Supplementary Figure 2**). We also evaluated age-adjusted excess mortality at the municipal level to obtain a more detailed overview and confirmed that excess mortality had a heterogeneous distribution at the municipal level, which was attributable to COVID-19 and non-COVID-19 causes (**Supplementary Figure 3**). Finally, at the state level, we observed that for non-COVID-19 deaths the highest decrease of in-hospital deaths was registered in Oaxaca, Yucatán, and Veracruz, whereas the highest proportion of non-COVID-19 out-of-hospital deaths were observed in Tlaxcala, Yucatan, and Colima (**Supplementary Figure 4**).

**Figure 3:**
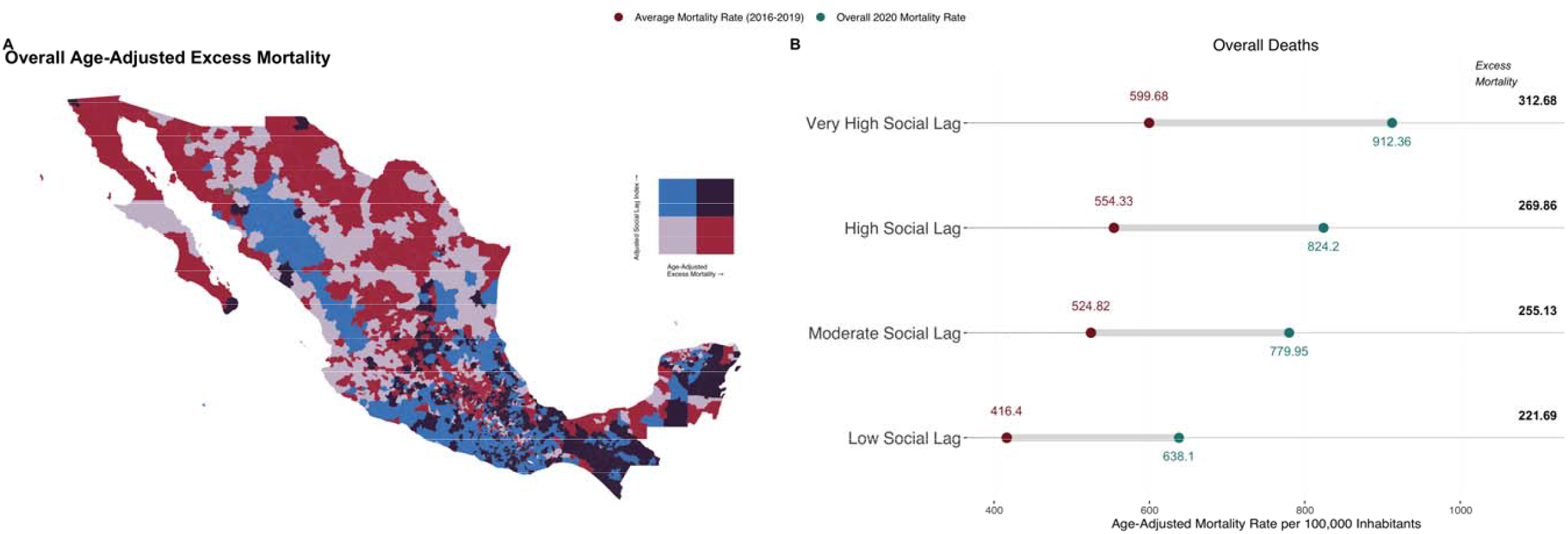
Choropleth map to visualize the geographical distribution of age-adjusted excess mortality and adjusted social lag index by municipality of occurrence in 2020 at Mexico (A). Historical and 2020 age-adjusted mortality rates and excess mortality stratified by adjusted social lag categories (B).

**Figure 4:**
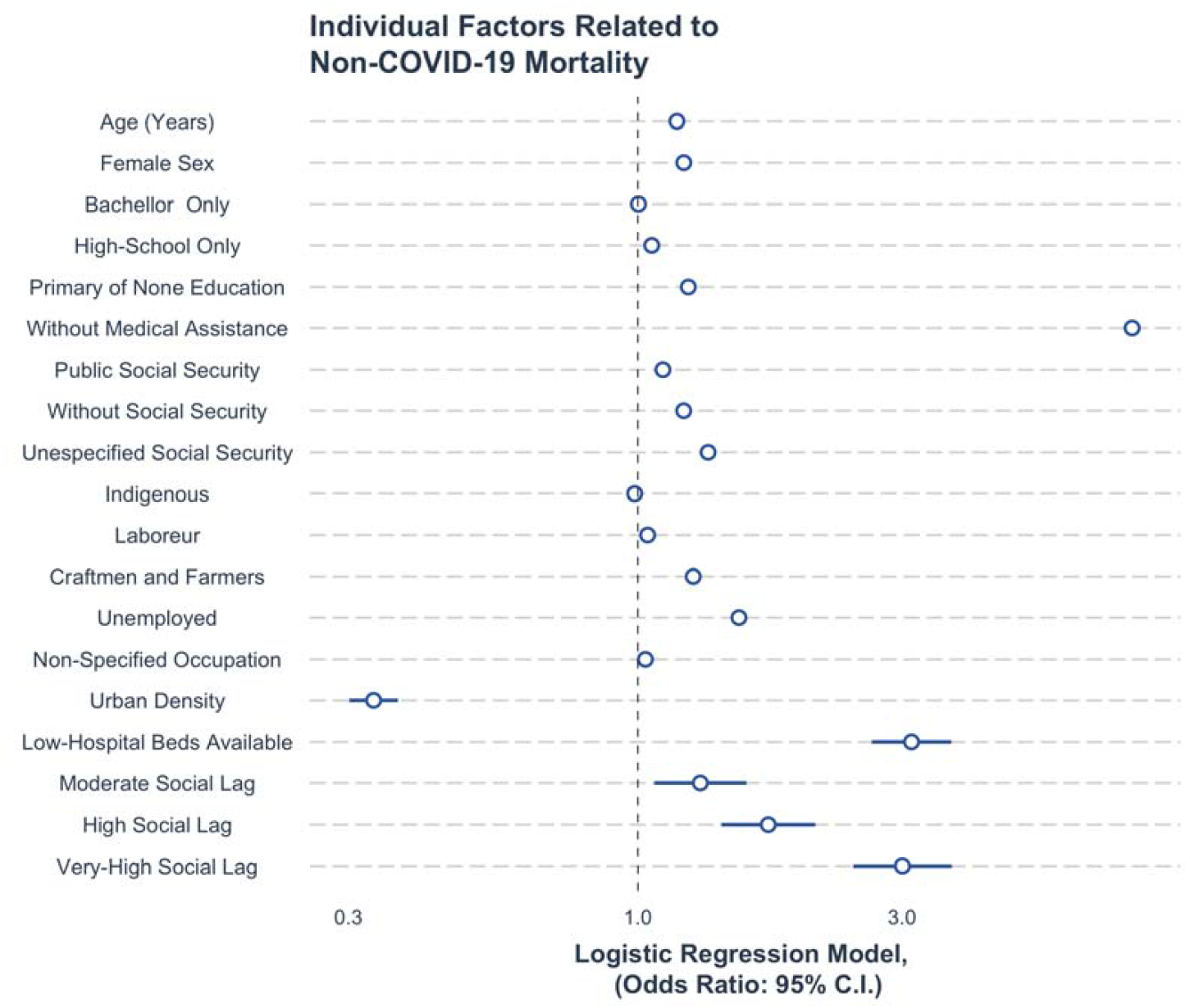
Binomial regression model to evaluate the municipal related factors associated with excess mortality in Mexico (A). Logistic regression model to evaluate individual related factors linked to the probability of death for non-COVID-19 causes (B).

### Municipal level impact of sociodemographic inequalities in excess mortality

We observed marked geographic variability in age-adjusted excess mortality across municipalities with higher aSLI (**Figure 3A**). Mexico’s southern and central municipalities were characterized by higher marginalization and excess mortality during 2020. After excluding COVID-19 related deaths, only the southern municipalities displayed the highest combination of excess mortality and aSLI (**Supplementary Figure 5**). Stratifying by aSLI categories, we observed that age-adjusted mortality rates and excess mortality displayed a stepwise increase with each higher marginalization level (**Figure 3B**). Municipalities classified with very high aSLI displayed both the higher age-adjusted mortality (921.36 per 100,000 inhabitants) and excess mortality (312.68 per 100,000 inhabitants) rates in Mexico. To evaluate the hypothesis that excess mortality was driven by social inequalities in healthcare access and hospital occupancy due to COVID-19 at the municipal level, we fitted negative binomial regression models for age-adjusted excess mortality rates. As previously observed, municipalities at high and very high social lag had the highest risk for non-COVID-19 age-adjusted excess mortality in 2020. Furthermore, municipalities with a higher percentage of population without social security coverage (IRR 1.03, 95%CI 1.02-1.04) and higher COVID-19 hospital occupancy (IRR 1.03, 95%CI 1.01-1.06) were at higher risk for excess mortality after adjusting for sex, education, medical assistance, urbanity, and death setting. We observed an interaction effect for higher risk for non-COVID-19 age-adjusted excess mortality in municipalities with very high social lag and higher COVID-19 hospital occupancy (IRR 1.08, 95%CI 1.03-1.12, **Supplementary Material**).

### Individual-level correlates of non-COVID 19 deaths

Finally, we explored the role of sociodemographic conditions and healthcare-related inequalities at an individual level for the risk for non-COVID 19 deaths using random-effects logistic regression models. We observed that women, people who had lower educational attainment and who worked as craftsmen, farmers, laborers, or were unemployed had an increased risk for death attributable to non-COVID-19 compared to COVID-19 causes. Regarding healthcare-related factors, we observed that people without medical assistance before death, people who reported a public or unspecified social security coverage or lived in municipalities with low availability of hospital beds had an increased risk of death for non-COVID-19 compared to COVID-19 causes. Finally, we confirm that people living in municipalities with high and very-high social lag had the highest risk for non-COVID-19 compared to COVID-19 death (**Figure 4**).

## DISCUSSION

In this study of 1,069,174 deaths recorded in Mexico between 2016-2020, we report that 49% of deaths in 2020 were in excess compared to the average reported between 2016-2019. Although cause-specific excess mortality during 2020 was largely attributable to COVID-19 (76.1%), non-COVID-19 causes comprised up to one-fifth of excess deaths in Mexico during 2020. Moreover, we report a differential impact in excess mortality related to the setting in which the deaths occurred; while COVID-19 deaths occurred primarily in-hospital, non-COVID-19 deaths sharply decreased in this setting and had a concurrent increase in the out-of-hospital setting. These findings contribute to the growing literature on the far-reaching impact of the COVID-19 pandemic on the health system and suggest both an excess in non-COVID-19 mortality as well as a displacement of these deaths to the out-of-hospital setting in Mexico.

We also observed that excess mortality displayed geographical heterogeneity linked to sociodemographic inequalities; states in the southern region of Mexico had the highest social marginalization and similarly high rates of non-COVID-19 excess mortality. We showed that lower prevalence of population without social security coverage and higher rates of COVID-19 hospitalization, combined with social marginalization, were municipal-level predictors of non-COVID-19 excess mortality. Finally, at the individual level, lower educational attainment, blue collar workers (laborers, craftsmen, and farmers), unemployment and lack of medical assistance prior deaths were significant predictors of non-COVID-19 compared to COVID-19 mortality during 2020. Overall, our results show that excess non-COVID-19 deaths could be assessed as a proxy of the failure of the healthcare system to provide adequate access to care for chronic diseases in situations of high demand and stress of an already fragile healthcare system (5). This situation is applicable to Mexico, but also to countries with similar socio-demographic profiles in the region or with increased rates of SARS-CoV-2 infections.

Previous reports have documented the high burden of excess mortality caused by the COVID-19 pandemic in Mexico, with excess mortality rates being estimated from 26.1 to 36.0 deaths per 100,000 inhabitants (6,13,17–19); moreover, Karlinsky et al. projected that Mexico’s actual toll of deaths could be twice the number of deaths registered during 2020 (20). These reports positioned Mexico as one of the leading countries in terms of excess mortality in Latin America and worldwide (6,20). However, there is limited information regarding cause-specific contributors to global excess mortality rates in Mexico. Our results demonstrate that COVID-19 deaths were significant contributors to excess mortality; nevertheless, we observed that excess deaths were also related to cardiometabolic chronic health conditions, including type 2 diabetes, cardiovascular disease, arterial hypertension, and obesity, which had a steep increase primarily in out-of-hospital settings. Excess non-COVID-19 deaths could be attributable to hospital reconversion policies and healthcare restructuration designed to improve care for COVID-19 cases, with the negative externality of reducing access to care for people with chronic health conditions who required continuous medical assistance during the COVID-19 pandemic. Other high-income countries have demonstrated the association between hospital occupancy and excess mortality during periods of peak COVID-19 infections (21–23). Explanations related to this phenomenon rely on data on restricted access to healthcare services in places which experienced hospital overload due to COVID-19, reduced out-of-hospital attention due to severely restricted healthcare services and personnel availability, lower insurance coverage, and lower number of healthcare personnel per-capita (24–26). Other reported non-related healthcare contributors were social stigma for being treated in hospitals due to potentially acquiring COVID-19 infection and increased unhealthy lifestyle habits which could have exacerbated complications due to chronic health conditions (27). Overall, excess non-COVID-19 mortality could be interpreted as an indirect indicator of the negative effects attributable to healthcare policies which prioritized in-hospital COVID-19 attention over the care of other chronic health conditions.

Notably, increased rates of COVID-19 hospitalizations and mortality in Mexico were observed in municipalities with high marginalization independently of urbanization, leading to increased stress in their healthcare systems (28,29); this phenomenon may explain the disproportionate impact on non-COVID-19 deaths in marginalized municipalities in Mexico. Our results support the hypothesis that groups with sociodemographic vulnerabilities experienced the highest impact of excess mortality attributable to hospital saturation, with this impact having an unequal distribution within Mexico. In the European region, countries with high excess mortality, such as Bulgaria, Russia, and Serbia were impacted by diverse social barriers with difficulties to full adhere to social isolation policies (30,31). Two recent reports in England revealed that communities with high-density of care homes, with a high proportion of residents on income support, overcrowding conditions, and ethnic minorities were at higher risk of excess mortality and years of life lost due to the COVID-19 pandemic (32,33). In Latin American countries, the impact of socioeconomic disparities was sharp mainly due to countries experiencing diverse social barriers to sustain lockdown mandates driven by low stipend support, a high proportion of their population working in informal conditions, and uncovered health care basic needs even within healthcare workers personnel (3,30,34). Nevertheless, this evidence and the comparison between countries should be interpreted with caution given the variation in COVID-19 dynamics, within-country gradients of sociodemographic inequalities, and the profiles of high-risk comorbidities.

Our results highlight the impact of healthcare-related and individual-level social inequalities in exacerbating overall and cause-specific excess mortality in Mexico. We show that the main contributor to higher non-COVID-19 excess mortality rates at the municipal level was a lower percentage of the population with access to social security health coverage. Furthermore, the interaction between a lower percentage of the population with access to social security health coverage and social marginalization confirmed the hazardous interplay between social and healthcare inequalities (35). At the individual level, we confirmed that certain socially vulnerable occupations experienced unequal risks for non-COVID-19 mortality. The role of individual and sociodemographic determinants in the risk for adverse COVID-19 outcomes has been previously explored (36,37). Nevertheless, our findings demonstrate that sociodemographic inequalities impacted individuals with preventable chronic conditions, regardless of public healthcare policies aimed at mitigating the impact of the COVID-19 pandemic in healthcare infrastructure and provision. Overall, our results represent an urgent call to action for local authorities to perform a healthcare restructuration, particularly in marginalized municipalities and with special attention for vulnerable groups to prioritize full coverage of hospital bed capacity, well-trained healthcare personnel, and availability of primary care services which cover chronic health conditions to prevent associated complications in the context of future COVID-19 waves or situations which increase stress and reduce access to healthcare in Mexico and other LMICs.

Our study has some strengths and limitations. Amongst the strengths, we highlight the use of 1,069,174 nationwide mortality registries to compare all-cause and cause-specific excess mortality during the COVID-19 pandemic in Mexico in 2020. This approach allowed us to estimate with higher confidence state and municipal-level excess morality rates that helped us to study the regional impact of the COVID-19 pandemic and identify vulnerable zones in Mexico which were especially affected during 2020 compared to previous years. Additionally, the use of sociodemographic variables at different levels gave us insights to evaluate municipal and individual-level determinants which shaped excess and non-COVID-19 mortality. Nevertheless, limitations to be acknowledged include the lack of specific clinical information and comorbidity assessment for predictors which have been proven to be crucial determinants that increase the risk of death for COVID-19 and non-COVID-19 causes, particularly regarding control of chronic cardio-metabolic conditions. Second, we could not ascertain the number of non-COVID-19 deaths which occurred due to exacerbation of underlying chronic conditions by current or previous SARS-CoV-2 infection, as it has been proven that it could increase the risk of long-term complications, including cardiovascular diseases (3). Third, our COVID-19 death construct included cases which could have been misclassified by atypical pneumonia or severe acute respiratory infections of unknown etiology, registered after the onset of the COVID-19 pandemic; this was done to reduce the risk of underreporting or misclassified COVID-19 deaths, but which could lead to overestimation of COVID-19 deaths. Finally, we used a surveillance dataset to assess COVID-19 hospitalization as a proxy to hospital occupancy; however, identification of COVID-19 related hospitalizations may have varied according to weekly SARS-CoV-2 testing capacity and adequate reporting. Therefore, the use of this proxy could be biased in municipalities with higher marginalization and reduced access to testing.

In conclusion, we show a high burden of excess mortality driven by in-hospital COVID-19 and out-of-hospital non-COVID-19 deaths. We observed regional heterogeneity of non-COVID-19 excess mortality, which was concentrated in marginalized municipalities in southern Mexico. High hospital occupancy due to COVID-19 and higher percentage of population without social security coverage were municipal wide-level determinants of excess mortality; whilst lower educational attainment, vulnerable working occupations, lack of medical assistance before death and being attended in public or underspecified health sectors were factors related to non-COVID-19 mortality likelihood. Our results demonstrate the impact of sociodemographic inequalities across all-levels in mortality related to non-COVID-19 causes in Mexico during 2020 compared to the 2016-2019 period. This data should prompt an urgent call to action to improve healthcare coverage and access, particularly in primary care settings to reduce health disparities in Mexico in situations which increase the stress of healthcare systems, including the ongoing COVID-19 pandemic.

## AUTHOR CONTRIBUTIONS

Research idea and study design: NEAV, CAFM, OYBC; data acquisition: NEAV, CAFM; data analysis/interpretation: NEAV, CAFM, JMA, LFC, JPA, AGD, AVV; statistical analysis: NEAV, CAFM, OYBC; manuscript drafting: NEAV, OYBC, CAFM, JMA, LFC, JPA, AGD, AVV, JAS SB, LMGR; supervision or mentorship: NEAV, OYBC. Each author contributed important intellectual content during manuscript drafting or revision and accepted accountability for the overall work by ensuring that questions pertaining to the accuracy or integrity of any portion of the work are appropriately investigated and resolved.

### FUNDING

This research was supported by Instituto Nacional de Geriatría.

## Data Availability

All code, datasets and materials are available for reproducibility of results at https://github.com/oyaxbell/excess_non_covid/

## ACKNOWLEDGMENTS

This project was registered and approved by the Research Committee at Instituto Nacional de Geriatría, project number DI-PI-006/2020. NEAV and CAFM are enrolled at the PECEM program of the Faculty of Medicine at UNAM. NEAV is supported by CONACyT. The authors would like to acknowledge the invaluable work of all of Mexico’s healthcare community in managing the COVID-19 pandemic. Their participation in the COVID-19 surveillance program along with open data sharing has made this work a reality, we are thankful for your effort.

## REFERENCES

1. Wang H, Paulson KR, Pease SA, Watson S, Comfort H, Zheng P, et al. Estimating excess mortality due to the COVID-19 pandemic: a systematic analysis of COVID-19-related mortality, 2020–21. Lancet [Internet]. 2022 Apr 16;399(10334):1513–36. Available from: https://doi.org/10.1016/S0140-6736(21)02796-3

2. Doubova S V, Leslie HH, Kruk ME, Pérez-Cuevas R, Arsenault C. Disruption in essential health services in Mexico during COVID-19: an interrupted time series analysis of health information system data. BMJ Glob Heal [Internet]. 2021 Sep 1;6(9):e006204. Available from: http://gh.bmj.com/content/6/9/e006204.abstract

3. Arceo-Gomez EO, Campos-Vazquez RM, Esquivel G, Alcaraz E, Martinez LA, Lopez NG. The income gradient in COVID-19 mortality and hospitalisation: An observational study with social security administrative records in Mexico. Lancet Reg Heal - Am [Internet]. 2022;6:100115. Available from: https://www.sciencedirect.com/science/article/pii/S2667193X21001113

4. Islam N. “Excess deaths” is the best metric for tracking the pandemic. BMJ [Internet]. 2022 Feb 4;376:o285. Available from: http://www.bmj.com/content/376/bmj.o285.abstract

5. Doubova S V, García-Saiso S, Pérez-Cuevas R, Sarabia-González O, Pacheco-Estrello P, Infante-Castañeda C, et al. Quality governance in a pluralistic health system: Mexican experience and challenges. Lancet Glob Heal [Internet]. 2018 Nov 1;6(11):e1149–52. Available from: https://doi.org/10.1016/S2214-109X(18)30321-8

6. Lima EEC, Vilela EA, Peralta A, Rocha M, Queiroz BL, Gonzaga MR, et al. Investigating regional excess mortality during 2020 COVID-19 pandemic in selected Latin American countries. Genus [Internet]. 2021;77(1):30. Available from: https://doi.org/10.1186/s41118-021-00139-1

7. Bello-Chavolla OY, Bahena-López JP, Antonio-Villa NE, Vargas-Vázquez A, González-Díaz A, Márquez-Salinas A, et al. Predicting mortality due to SARS-CoV-2: A mechanistic score relating obesity and diabetes to COVID–19 outcomes in Mexico. J Clin Endocrinol Metab [Internet]. 2020 May 31; Available from: https://doi.org/10.1210/clinem/dgaa346

8. Gutiérrez JP, García-Saisó S. Health inequalities: Mexico’s greatest challenge. Lancet [Internet]. 2016 Nov 12;388(10058):2330–1. Available from: https://doi.org/10.1016/S0140-6736(16)31726-3

9. Palacio-Mejía LS, Hernandez-Avila JE, Hernandez-Avila M, Dyer Leal D, Barranco Flores A, Quezada Sanchez AD, et al. Leading Causes of Excess Mortality in Mexico During the COVID-19 Pandemic 2020 – February 2021: A Death Certificates Study in a Lower-Middle-Income Country. Rochester, NY: Social Science Research Network; 2021 Apr.

10. Instituto Nacional de Estadística y Geografía. Datos de Mortalidad en México [Internet]. 2021 [cited 2021 Mar 1]. Available from: https://www.inegi.org.mx/programas/mortalidad/#Documentacion

11. Kontopantelis E, Mamas MA, Webb RT, Castro A, Rutter MK, Gale CP, et al. Excess deaths from COVID-19 and other causes by region, neighbourhood deprivation level and place of death during the first 30 weeks of the pandemic in England and Wales: A retrospective registry study. Lancet Reg Heal – Eur [Internet]. 2021 Aug 1;7. Available from: https://doi.org/10.1016/j.lanepe.2021.100144

12. Dirección General de Información en Salud. Catálogos de Defunciones. 2021.

13. Karlinsky A, Kobak D. Tracking excess mortality across countries during the COVID-19 pandemic with the World Mortality Dataset. Davenport MP, Lipsitch M, Lipsitch M, Simonsen L, Mahmud A, editors. Elife [Internet]. 2021;10:e69336. Available from: https://doi.org/10.7554/eLife.69336

14. CONEVAL. Índice Rezago Social [Internet]. 2015. Available from: https://www.coneval.org.mx/Medicion/IRS/Paginas/Indice_Rezago_Social_2015.aspx

15. Direccion General de Epidemiología. COVID-19/MEXICO [Internet]. 2020 [cited 2020 Dec 28]. Available from: https://datos.covid-19.conacyt.mx

16. INEGI. Metodología de Indicadores de la Serie Histórica Censal [Internet]. 2018 [cited 2021 Jan 31]. Available from: https://inegi.org.mx/contenidos/programas/ccpv/cpvsh/doc/serie_historica_censal_met_indicadores.pdf

17. Palacio-Mejía LS, Wheatley-Fernández JL, Ordóñez-Hernández. All-cause excess mortality during the Covid-19 pandemic in Mexico. Salud Publica Mex. 2021;63(2):211–24.

18. Dahal S, Banda JM, Bento AI, Mizumoto K, Chowell G. Characterizing all-cause excess mortality patterns during COVID-19 pandemic in Mexico. BMC Infect Dis [Internet]. 2021;21(1):432. Available from: https://doi.org/10.1186/s12879-021-06122-7

19. Dahal S, Luo R, Swahn MH, Chowell G. Geospatial Variability in Excess Death Rates during the COVID-19 Pandemic in Mexico: Examining Socio Demographic, Climate and Population Health Characteristics. Int J Infect Dis [Internet]. 2021;113:347–54. Available from: https://www.sciencedirect.com/science/article/pii/S1201971221008146

20. Karlinsky A, Kobak D. The World Mortality Dataset: Tracking excess mortality across countries during the COVID-19 pandemic. medRxiv Prepr Serv Heal Sci [Internet]. 2021 Jun 4;2021.01.27.21250604. Available from: https://pubmed.ncbi.nlm.nih.gov/33532789

21. French G, Hulse M, Nguyen D, Sobotka K, Webster K, Corman J, et al. Impact of hospital strain on excess deaths during the COVID-19 pandemic-United States, july 2020-july 2021. Am J Transplant Off J Am Soc Transplant Am Soc Transpl Surg. 2022 Feb;22(2):654–7.

22. Michelozzi P, de’Donato F, Scortichini M, Pezzotti P, Stafoggia M, De Sario M, et al. Temporal dynamics in total excess mortality and COVID-19 deaths in Italian cities. BMC Public Health [Internet]. 2020;20(1):1238. Available from: https://doi.org/10.1186/s12889-020-09335-8

23. Rossman H, Meir T, Somer J, Shilo S, Gutman R, Ben Arie A, et al. Hospital load and increased COVID-19 related mortality in Israel. Nat Commun [Internet]. 2021;12(1):1904. Available from: https://doi.org/10.1038/s41467-021-22214-z

24. Pilkington H, Feuillet T, Rican S, de Bouillé JG, Bouchaud O, Cailhol J, et al. Spatial determinants of excess all-cause mortality during the first wave of the COVID-19 epidemic in France. BMC Public Health. 2021 Nov;21(1):2157.

25. Stokes AC, Lundberg DJ, Bor J, Elo IT, Hempstead K, Preston SH. Association of Health Care Factors With Excess Deaths Not Assigned to COVID-19 in the US. JAMA Netw open. 2021 Sep;4(9):e2125287.

26. Watkins DA. Cardiovascular health and COVID-19: time to reinvent our systems and rethink our research priorities. Heart [Internet]. 2020 Dec 1;106(24):1870 LP –1872. Available from: http://heart.bmj.com/content/106/24/1870.abstract

27. Bagcchi S. Stigma during the COVID-19 pandemic. Lancet Infect Dis [Internet]. 2020 Jul;20(7):782. Available from: https://pubmed.ncbi.nlm.nih.gov/32592670

28. Naja F, Hamadeh R. Nutrition amid the COVID-19 pandemic: a multi-level framework for action. Eur J Clin Nutr [Internet]. 2020;74(8):1117–21. Available from: https://doi.org/10.1038/s41430-020-0634-3

29. Zhang J. Hospital Avoidance and Unintended Deaths during the COVID-19 Pandemic. Am J Heal Econ [Internet]. 2021 May 4;7(4):405–26. Available from: https://doi.org/10.1086/715158

30. Kapitsinis N. The underlying factors of excess mortality in 2020: a cross-country analysis of pre-pandemic healthcare conditions and strategies to cope with Covid-19. BMC Health Serv Res [Internet]. 2021;21(1):1197. Available from: https://doi.org/10.1186/s12913-021-07169-7

31. Gallo V, Mackenbach JP, Ezzati M, Menvielle G, Kunst AE, Rohrmann S, et al. Social Inequalities and Mortality in Europe – Results from a Large Multi-National Cohort. PLoS One [Internet]. 2012 Jul 25;7(7):e39013. Available from: https://doi.org/10.1371/journal.pone.0039013

32. Davies B, Parkes BL, Bennett J, Fecht D, Blangiardo M, Ezzati M, et al. Community factors and excess mortality in first wave of the COVID-19 pandemic in England. Nat Commun [Internet]. 2021;12(1):3755. Available from: https://doi.org/10.1038/s41467-021-23935-x

33. Kontopantelis E, Mamas MA, Webb RT, Castro A, Rutter MK, Gale CP, et al. Excess years of life lost to COVID-19 and other causes of death by sex, neighbourhood deprivation, and region in England and Wales during 2020: A registry-based study. PLOS Med [Internet]. 2022 Feb 15;19(2):e1003904. Available from: https://doi.org/10.1371/journal.pmed.1003904

34. Antonio-Villa NE, Bello-Chavolla OY, Vargas-Vázquez A, Fermín-Martínez CA, Márquez-Salinas A, Pisanty-Alatorre J, et al. Assessing the Burden of Coronavirus Disease 2019 (COVID-19) Among Healthcare Workers in Mexico City: A Data-Driven Call to Action. Clin Infect Dis an Off Publ Infect Dis Soc Am. 2021 Jul;73(1):e191–8.

35. Ji Y, Ma Z, Peppelenbosch MP, Pan Q. Potential association between COVID-19 mortality and health-care resource availability. Vol. 8, The Lancet. Global health. 2020. p. e480.

36. Fielding-Miller RK, Sundaram ME, Brouwer K. Social determinants of COVID-19 mortality at the county level. medRxiv Prepr Serv Heal Sci [Internet]. 2020 Jul 1;2020.05.03.20089698. Available from: https://pubmed.ncbi.nlm.nih.gov/32637976

37. Antonio-Villa NE, Fernandez-Chirino L, Pisanty-Alatorre J, Mancilla-Galindo J, Kammar-García A, Vargas-Vázquez A, et al. Comprehensive Evaluation of the Impact of Sociodemographic Inequalities on Adverse Outcomes and Excess Mortality During the Coronavirus Disease 2019 (COVID-19) Pandemic in Mexico City. Clin Infect Dis [Internet]. 2021 Jun 22;ciab577. Available from: https://doi.org/10.1093/cid/ciab577

