## Supplementary Material for "Sociodemographic inequalities and excess non-COVID-19 mortality during the COVID-19 pandemic: A data-driven analysis of 1,069,174 death certificates in Mexico"

**Death certification and validation from National Institute of Statistics and Geography (INEGI) dataset**

INEGI takes as a unit of measurement, the death certificate registered in all health units and hospitals registered in Mexico. The death certificate is the main capture format to generate mortality statistics, and which is issued by medical professionals or persons authorized by the competent health authority, once the death and its causes have been verified, in the approved models by the Secretary of Health and in accordance with the technical standards that it issues. The death certificate has one original and three copies, the original remaining for the Ministry of Health, a copy is distributed to INEGI, a copy for the Civil Registry and as of 2010 a copy is attached for the medical unit that certifies the death. The death certificate has one original and three copies, the original remaining for the Ministry of Health, a copy is distributed to INEGI, a copy for the Civil Registry and as of 2010 a copy is attached for the medical unit that certifies the death.

**National Epidemiological Surveillance Study (SINAVE) dataset**

The database holds information on all persons tested for SARS-CoV-2 infection at public facilities in Mexico, as well as in all private healthcare facilities that follow the legal mandate to report COVID-19 cases to health authorities and the public locations for rapid antigen testing approved by the SS. All demographic and health data were collected and uploaded to the NESS database by healthcare personnel from each corresponding healthcare facility. Available variables include age, sex, nationality, state, and municipality where the case was detected, immigration status as well as identification of individuals who self-identify as indigenous. Health information includes the status of diabetes, obesity, chronic obstructive pulmonary disease (COPD), immunosuppression, pregnancy, arterial hypertension, cardiovascular disease, chronic kidney disease (CKD), and asthma. Evaluated symptoms included fever, cough, odynophagia, dyspnea, irritability, diarrhea, chest pain, shivering, headache, myalgia, arthralgia, malaise, rhinorrhea, polypnea, vomiting, abdominal pain, conjunctivitis, cyanosis, and sudden onset of symptoms.

**Social lag index components**

The SLI is composite of several components that are measured to estimate social disadvantage and structural inequality at a municipal level based on population census data from the Mexican National Evaluation Council (CONEVAL); SLI is a principal component score which comprises percentages of literacy, access to basic education, healthcare services, living conditions including drainage, dirt floor, access to water, electricity, and electrical appliances for each Mexican municipality. Although CONEVAL proposes a categorization by quintiles to assess social lag by municipality for the entire country, Mexico City represents an exception. The municipalities of Mexico City are categorized as a low lag compared to the rest of the country.

**Supplementary Figure 1:** Age adjusted mortality rate for COVID-19 and non-COVID-19 deaths (A). Percentage of age-adjusted excess mortality rates attributable to COVID-19 and non-COVID-19 deaths (B).

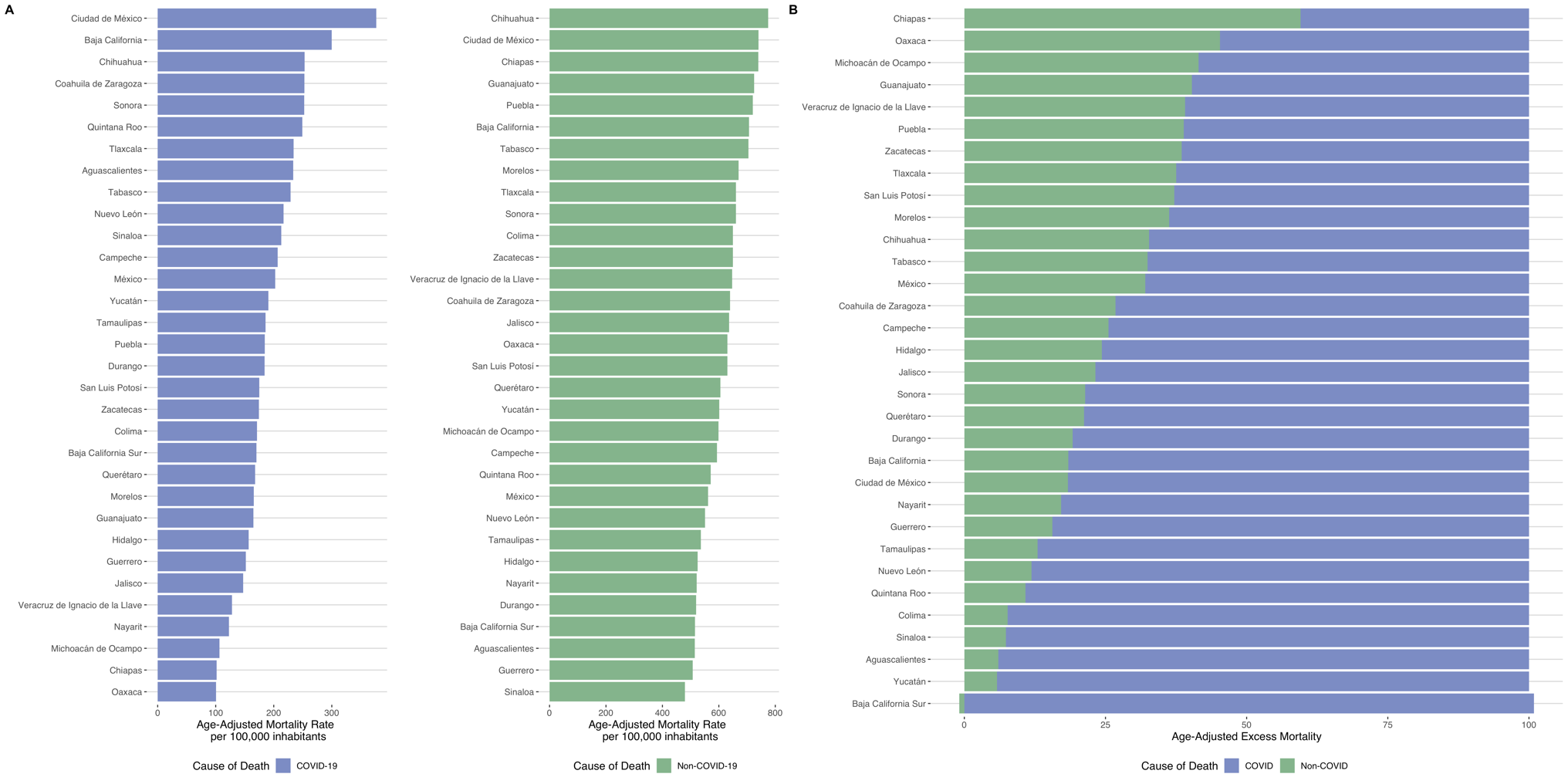

**Supplementary Figure 2:** Excess age-adjusted excess mortality for the main 10 causes of death in Mexico during the year 2020.
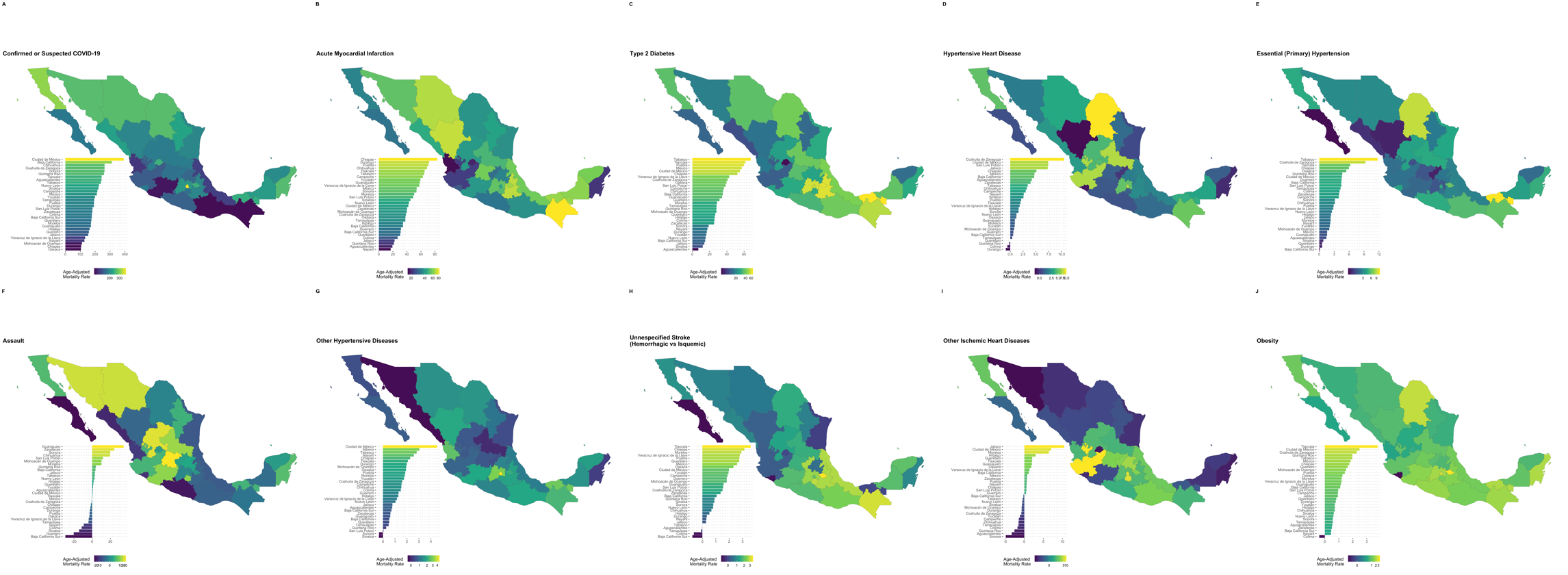

**Supplementary Figure 3:** Choropleth map by municipalities with higher percentage of age-adjusted excess mortality attributable to COVID-19 and non-COVID-19 causes in Mexico during 2020.

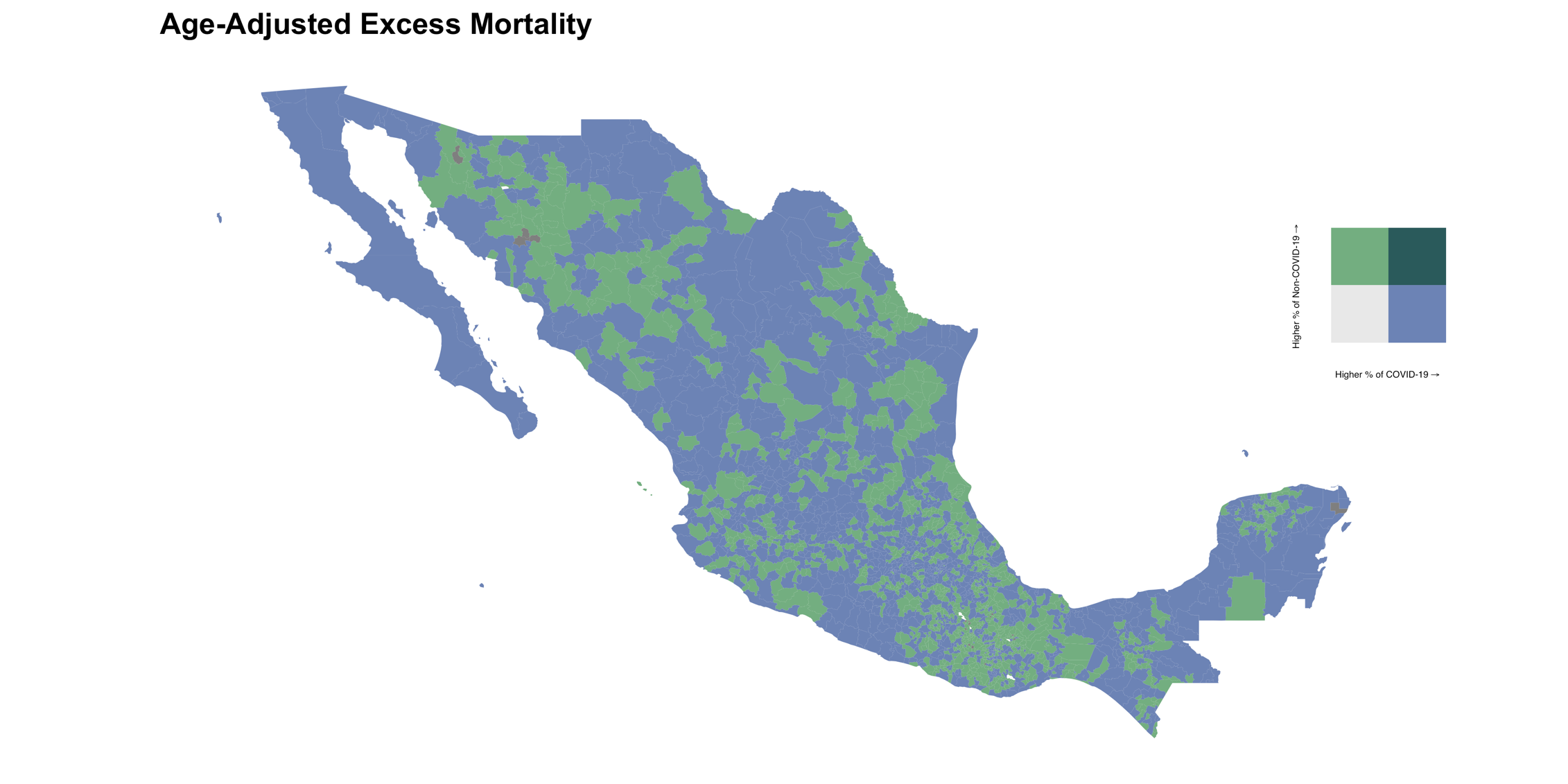

**Supplementary Figure 4:** Age adjusted mortality rate for COVID-19 and non-COVID-19 deaths stratified by death setting (A). Percentage of age-adjusted excess mortality rates attributable to COVID-19 and non-COVID-19 deaths stratified by death setting (B).

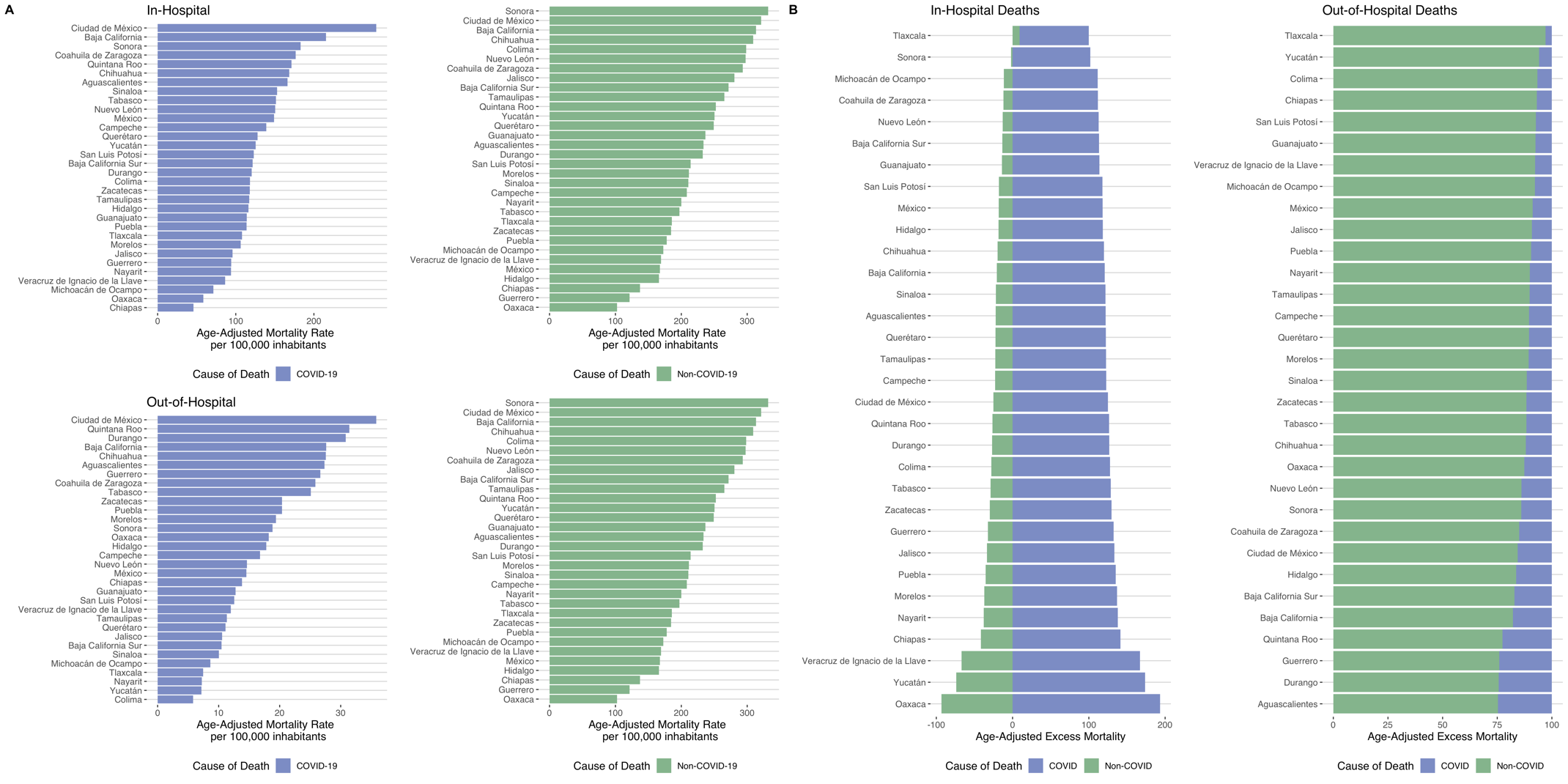

**Supplementary Figure 5:** Choropleth map by municipalities with higher age-adjusted excess mortality attributable to COVID-19 (A) and non-COVID-19 causes (B) related with adjusted social lag index in Mexico during 2020.

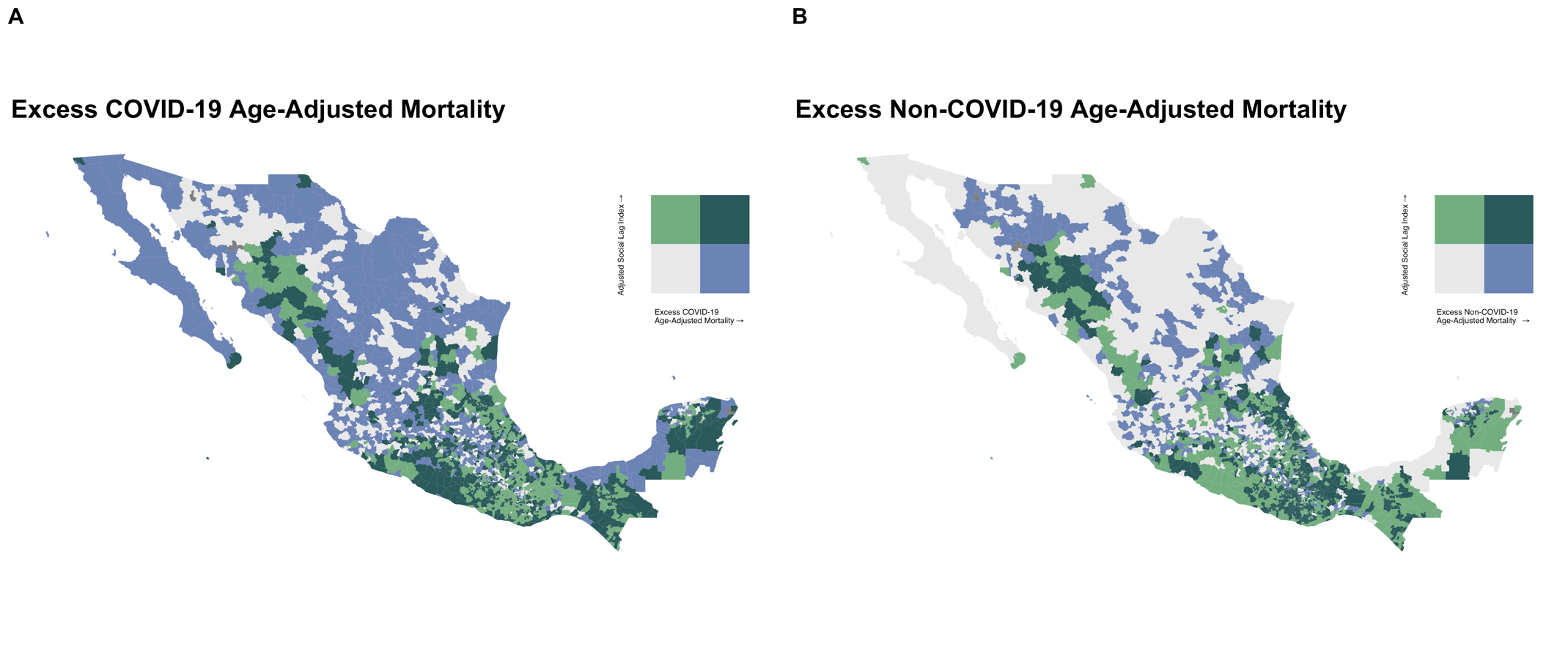

**Supplementary Figure 6:** Incidence Rate-Ratio plots for negative binomial logistic regression models to assess the interaction effect of higher COVID-19 hospitalization and percentage of inhabitants without social security and social lag categories.
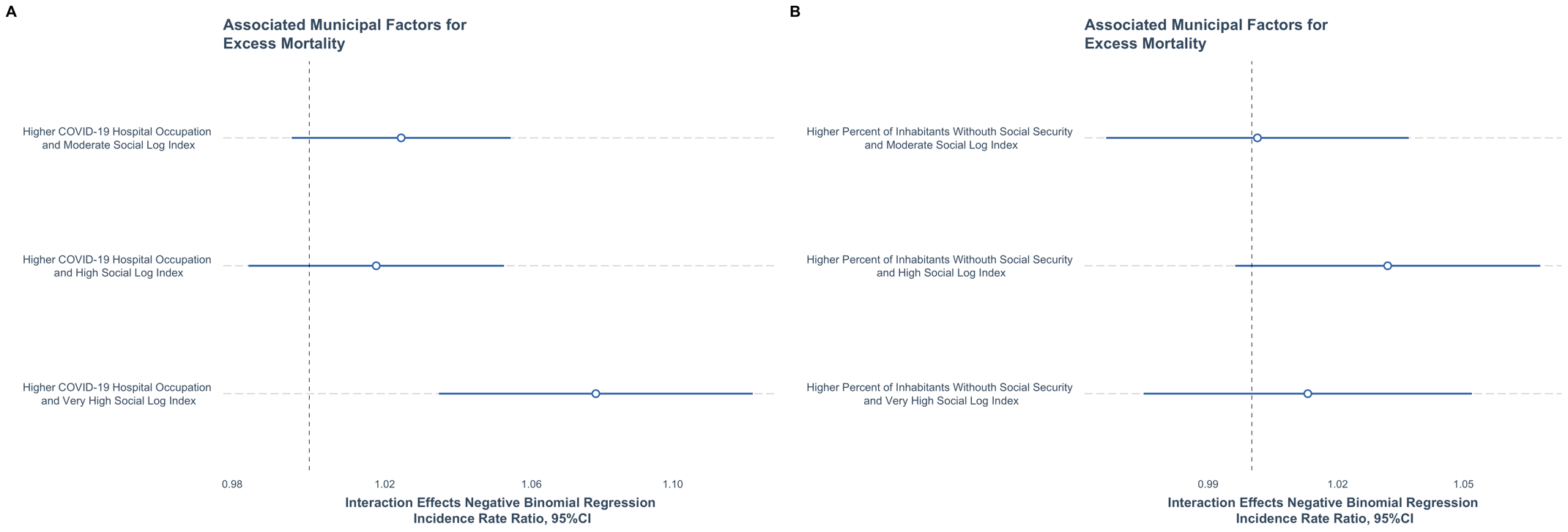

**Supplementary Table 2:** Estimated overall age-adjusted mortality and excess mortality stratified by death setting in Mexico for the year 2020.

| Death Setting | Deaths in 2020 | 2020 Mortality rate per 100,000 | COVID-19 Deaths per 100,000 | Non-COVID-19 deaths per 100,000 | Average deaths 2016-2019 | Average Mortality per 100,000 for 2016-2019 | Excess Mortality per 100,000 | % Attributable to COVID-19 deaths | % Attributable to Non-COVID-19 deaths |
| --- | --- | --- | --- | --- | --- | --- | --- | --- | --- |
| In-Hospital | 460,651 | 360.17 | 166.57 | 193.59 | 311,063 | 249.21 | 110.96 | 150.11% | - 50.12% |
| Out-of-Hospital | 583,597 | 455.56 | 30.93 | 424.62 | 372,086 | 297.45 | 158.10 | 19.56% | 80.43% |
| Unspecified | 24,926 | 17.76 | 1.74 | 16.01 | 14,931 | 10.72 | 7.04 | 24.79% | 75.20% |
